# Short-term versus 6-week prednisone in the treatment of subacute thyroiditis: a randomized controlled trial

**DOI:** 10.1101/2020.02.15.20023283

**Authors:** Lian Duan, Xiaoli Feng, Rui Zhang, Xiaojuan Tan, Xiaoyan Xiang, Rufei Shen, Hongting Zheng

**Author notes:** Correspondence: Dr. Lian Duan, ORCiD numbers: 0000-0003-4272-3461, The Third Affiliated Hospital of Chongqing Medical University (Jie er Hospital), Chongqing 401120, China. Author details: Lian Duan, Shuanghu branch No.1, Yubei District, Chongqing 401120, China. Xiaoli Feng, Xinqiao Street No.183, Shapingba District, Chongqing 400037, China. Rui Zhang, Xinqiao Street No.183, Shapingba District, Chongqing 400037, China. Xiaojuan Tan, Xinqiao Street No.183, Shapingba District, Chongqing 400037, China. Xiaoyan Xiang, Xinqiao Street No.183, Shapingba District, Chongqing 400037, China. Rufei Shen, Xinqiao Street No.183, Shapingba District, Chongqing 400037, China. Hongting Zheng, Xinqiao Street No.183, Shapingba District, Chongqing 400037, China.

## Abstract

**Background:** Moderate-to-severe subacute thyroiditis is commonly treated with 6-8 weeks glucocorticoids; however, no studies have described short-term prednisone treatment for subacute thyroiditis. We evaluated the efficacy of this treatment for subacute thyroiditis.

**Methods:** This was a 24-week, prospective, single-blind, randomized controlled study. Patients aged 18-70 years with subacute thyroiditis were hospitalized from August 2013 to December 2014. Patients with moderate-to-severe symptoms were randomized to receive either 30 mg/d prednisone for 1 week and then switched to 1 week of nonsteroidal anti-inflammatory drugs or 6 weeks of prednisone. The primary endpoints were the differences in efficacy at the end of treatment between two groups. Secondary endpoints included differences between the two groups in parameters of side effects at withdrawal and thyroid function at weeks 6, 12, 24.

**Results:** Of 96 patients screened, 52 subjects were randomized and 50 completed the study. Efficacy and recurrence rates were not significantly different at withdrawal in both groups (P=0.65). Parathyroid hormone (28.8 vs 38.9 pg/ml, p=0.011) and mean systolic blood pressure (113.9 vs 122.4 mmHg, p=0.023) were significantly lower in the experimental group than in the control group at discontinuation. No significant differences were observed in other secondary endpoints at withdrawal and in thyroid function at the 6th, 12th and 24th week during the follow-up time between the two groups.

**Conclusions:** Fewer side effects of glucocorticoids and similar efficacy and recurrence rates were observed with short-term prednisone compared with those with 6-week treatment for subacute thyroiditis. Short-term prednisone with a better safety profile may be as one alternative strategy for ameliorating moderate-to-severe symptoms of subacute thyroiditis.

**Trial registration:** Trial registration number NCT01837433. Registered with Clinicaltrials.gov on 23 April 2013

## Background

Subacute thyroiditis (SAT) is the most common painful thyroid disease and is a common cause of thyrotoxicosis. It is diagnosed based on the following: viral infection induction, fever, thyrotoxicosis, enlarged firm thyroid, palpation pain, radiation of pain to the parietal and occipital regions and to the ears, jaw, or throat, high erythrocyte sedimentation rate (ESR) and elevated C-reactive protein (CRP) levels. SAT can be accompanied by mild anemia, low thyroid iodine uptake rate, diffuse heterogeneous changes in ultrasonography, and decreased or normal color doppler blood flow signal [1]. The American Thyroid Association guidelines recommend glucocorticoid therapy (prednisone 40 mg daily for 1-2 weeks followed by a gradual taper over 2-4 weeks or longer, depending on clinical response) for patients in whom nonsteroidal anti-inflammatory drugs (NSAIDs) show poor efficacy or for those with moderate-to-severe symptoms at onset [2]. The Chinese guidelines recommend starting therapy with 20-40 mg/d prednisone for 1-2 weeks and then slowly reducing according to symptoms, signs and ESR. The usual course is 6-8 weeks. However, at least 20% of patients have a course of treatment longer than 8 weeks in one study [3]. at least 20% of patients have a course of treatment greater than eight weeks

However, long-term glucocorticoids can increase blood glucose, blood pressure, blood lipid levels, bone metabolism disorders [4], and cause Cushing’s features, chronic hypoadrenocorticism, and even acute adrenal crisis [5]. Therefore an ideal therapy would limit the duration of treatment. Thus far, no randomized controlled trials (RCTs) investigating short-term glucocorticoid treatment protocols are available. We therefore conducted a RCT to assess the efficacy and safety of short-term prednisone therapy for moderate-to-severe SAT.

## Methods

### Study design and participants

We conducted a 24-week prospective, single-blind, randomized controlled clinical trial at the department of endocrinology, the second affiliated hospital, third military medical university, Chongqing, China. This study was approved by the ethics committee of the second affiliated hospital, third military medical university (reference number 2018012-01). All patients signed written informed consent forms according to the tenets of the Declaration of Helsinki. SAT was diagnosed based on diagnostic criteria [2]. The severity of SAT was scored [6] as follows: fever – none 0 point, <38□ 1 point, and >38□ 2 points; tenderness – none 0 point, mild 1 point, and severe 2 points; goiter by ultrasonography or palpation – none 0 point and yes 1 point; and ESR – normal 0 point, 25-60 mm/h 1 point, and >60 mm/h 2 points. 18-70 years patients with SAT scored ≥ 3 points were included. Patients who were diagnosed with diabetes, an active peptic ulcer, a tumor, hepatic dysfunction, recurrent SAT and those who were already taking glucocorticoids were excluded.

Moderate-to-severe SAT was defined by us as a definite diagnosis of SAT, with a sore thyroid, and a score ≥3 points. Efficacy was defined as no pain in the thyroid region, disappearance of clinical symptoms such as thyroid tenderness [7], and normalization of ESR and/or CRP level. Recurrence was defined as the development of a goiter with tenderness at the neck, fever, and re-elevation of inflammatory markers during prednisone tapering or after therapy withdrawal [8]. Anemia was defined as a hemoglobin (Hb) level <130 g/L for males and <120 g/L for females [9]. Overt and subclinical hyperthyroidism were defined as decreased serum thyroid-stimulating hormone (TSH) levels with elevated or normal levels of free triiodothyronine (FT3) and/or free thyroxine (FT4) (1). Subclinical hypothyroidism was characterized by a TSH level above the upper reference limit in combination with a normal FT4 level. An elevated TSH level (>10 mIU/L) in combination with a subnormal FT4 level characterized overt hypothyroidism [10]. Persistent hypothyroidism was defined by us as hypothyroidism that continued after the 24-week follow-up.

### Randomization and masking

Patients were randomly assigned to short-term or 6-week prednisone treatment (Fig. 1) in accordance with a random number and masked to randomization and treatment allocation.

**Figure 1.**
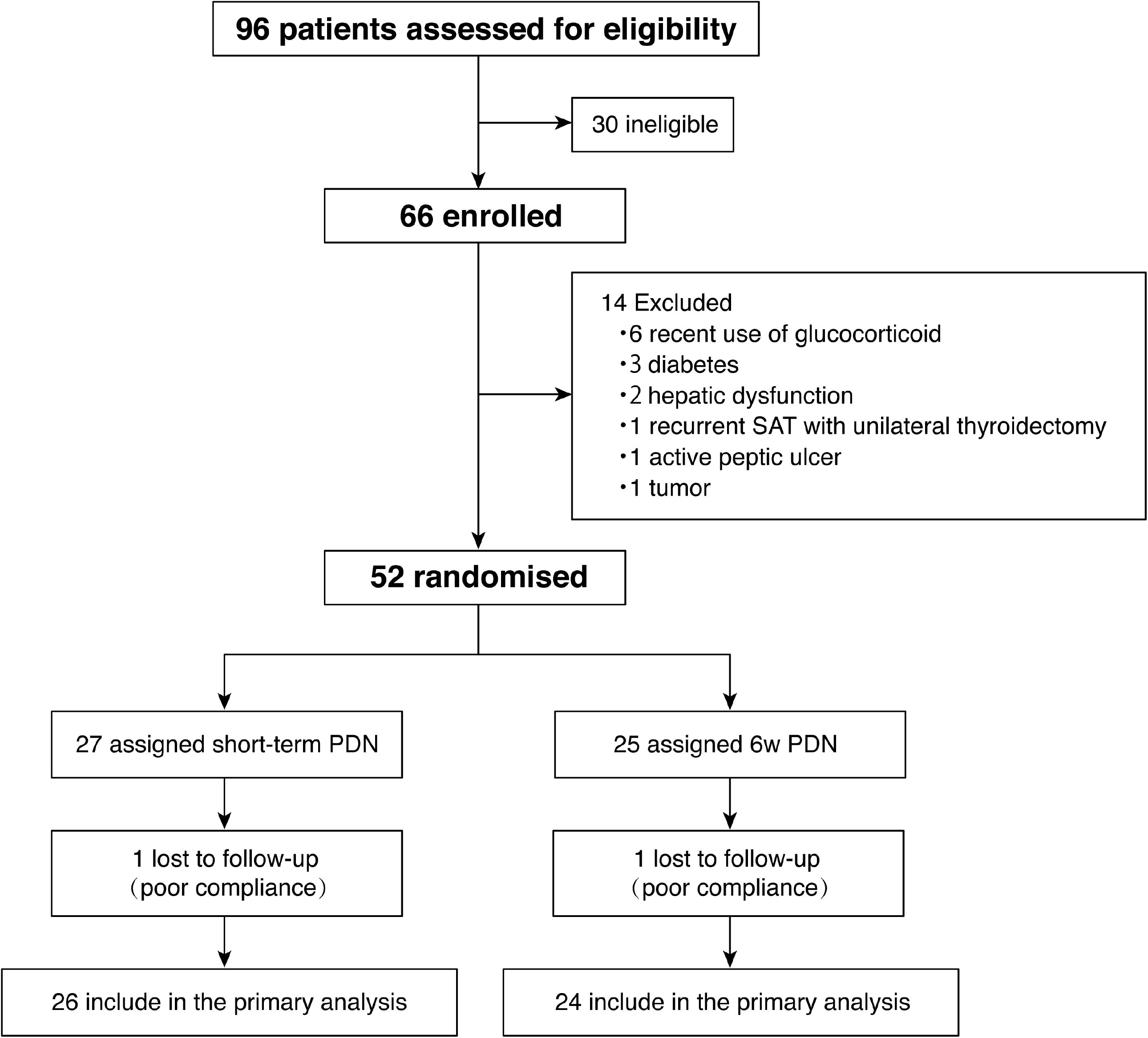
Trial profile PDN=Prednisone.

### Procedures

The patients with moderate-to-severe SAT were treated as follows: 1) those in the experimental group received 20 mg prednisone in the morning and 10 mg in the afternoon, daily for 1 week. In the 2nd week they received 400 mg celecoxib on the 1st day and 200 mg twice daily during the remaining 6 days until celecoxib withdrawal; and 2) those in the control group received 20 mg prednisone in the morning and 10 mg in the afternoon, daily in the 1st week. The dose was then reduced by 5 mg/week from the 2nd week until withdrawal in the 6th week. All patients were followed up for 24 weeks after initial treatment (Fig. 2). The experimental group had five followed-up visits. ESR, CRP and cortisol (8 am) were surveyed in the 1st and 2nd week after the initial treatment. Glycated albumin (GA), blood glucose, bone metabolism, blood lipids, and blood pressure were investigated in the 2nd week after the initial treatment. The control group had four follow-up visits. ESR, CRP and cortisol (8 am) were surveyed in the 1st week after the initial treatment. GA, blood glucose, bone metabolism, blood lipids, blood pressure and cortisol (8 am) were investigated in the 6^th^ week after the initial treatment. Thyroid function was measured in the 6th, 12th and 24th week after the initial treatment in both groups. Each group lost one patient during the follow-up because of poor compliance, resulting in a total of 50 patients completing the follow-up. Proton pump inhibitors were administered to all patients. Relapsed patients were administered prednisone 10 mg twice daily and tapered by 5 mg/week after symptom control for 4 weeks during treatment and after withdrawal.

**Figure 2.**
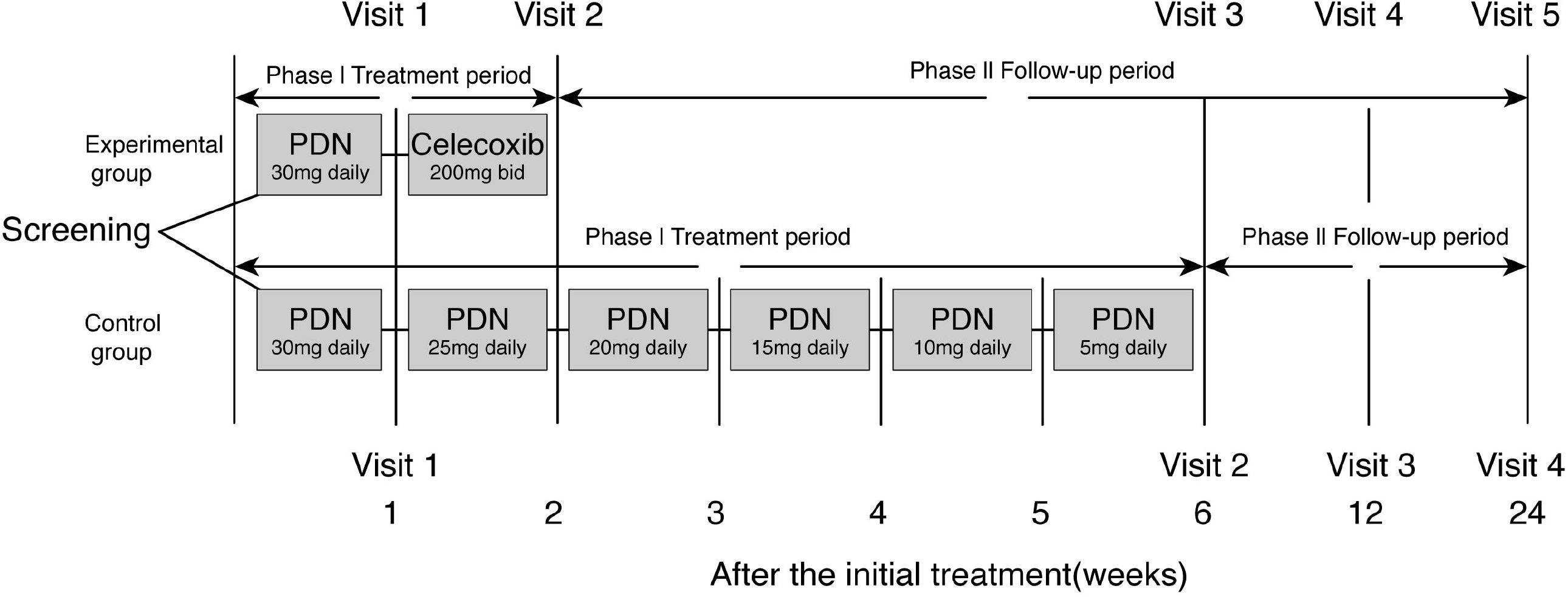
Study protocol PDN=Prednisone.bid=twice daily. W=week.

### Outcomes

The primary endpoints were the differences between the two groups in efficacy and recurrence rates at the end of treatment. The secondary endpoints included differences between the two groups in cortisol, total cholesterol (TC), triglyceride (TG) levels, systolic blood pressure (SBP), diastolic blood pressure (DBP), β cross-linked C-telopeptide of type 1 collagen (BCTX), osteocalcin (OC), type I procollagen amino terminal peptide (PINP), parathyroid hormone (PTH), 25-hydroxy vitamin D (25-OH-VD), calcitonin (CT), peripheral blood glucose, GA levels, and thyroid function [FT3, FT4, TSH, serum anti-thyroglobulin antibody (TGAb), serum anti-thyroid peroxidase antibody (TPOAb), thyroid-binding globulin (TBG), and serum anti-TSH receptor antibody (TRAb) levels].

### Statistical analysis

Based on past intervention trials [11-17], the sample numbers ranged from 5 to 49 cases. The approximate number of hospitalized patients with SAT in our department was 50 per year. Considering that 5% of patients may have been lost during follow-up, we decided to include 52 patients.

Data with a normal distribution were presented with means and standard deviations (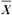 ± *s*), and data with a skewed distribution were presented with medians and interquartile ranges (P25-P75). Categorical data were presented by numbers and percent (%). Two independent sample t-tests and two independent sample Wilcoxon rank sum tests were performed to compare the difference between the two treatment groups for the continuous data with normal distributions and those with skewed distributions, respectively. Fisher’s exact test was performed to assess the associations of categorical data with treatment groups. The changes between two time points within the treatment groups were evaluated by a paired t-test, and the non-parametric Wilcoxon signed ranks test, respectively, for continuous data with normal distributions, and for the data with a skewed distributions. A generalized linear mixed model was used to test repeated measures of data due to missing values. Statistical analyses were evaluated at a two-sided significance level of 0.05 and were performed using SAS 9.4 software (SAS Institute Inc., Cary, NC, USA).

## Results

### Baselines

From August 1, 2013, to December 31, 2014, 96 hospitalized individuals were screened for eligibility. In total, 30 cases with mild symptoms whose scores were less than 3 points were ineligible. Furthermore, 14 cases were excluded because of recent use of glucocorticoids, diabetes mellitus, hepatic dysfunction, recurrent SAT with unilateral thyroidectomy, active peptic ulcer, and a tumor. Thus, 52 patients with moderate-to-severe SAT were randomized to the experimental and control groups in the study. Finally, data were obtained from 50 patients (Fig. 1). The two groups had comparable ages, male to female proportions, severity scores, radioiodine uptake (RAIU) at 3 and 24 h, Hb, TPOAb, TGAb, TBG, and TRAb levels (Table 1).

**Table 1.** Baseline characteristics

| Characteristic | Short-term<br>treatment<br>(n=26) | 6-weeks<br>treatment<br>(n=24) | Statistical<br>magnitude | P-value |
| --- | --- | --- | --- | --- |
| Age (year) | 45.54±8.87 | 44.08±9.51 | 0.56<sup>T</sup> | 0.578 |
| Gender | 20(76.92%) | 20(83.33%) | / | 0.728<sup>F</sup> |
| Severity score | 4(3-6) | 4(3-4.5) | -1.191<sup>Z</sup> | 0.233 |
| 3h RAIU (%) | 1.45(1.2-1.7) | 1.45(1.15-1.7) | -0.029<sup>Z</sup> | 0.977 |
| 24h RAIU (%) | 1.15(0.3-1.8) | 1.6(1.05-1.95) | 0.065<sup>Z</sup> | 0.129 |
| Hb (g/L) | 115.5(111.0-124.0) | 114.5(110-126.5) | 0.204<sup>Z</sup> | 0.838 |
| TPOAb | 10(5-24) | 17(6-40) | 0.969<sup>Z</sup> | 0.322 |
| TGAb (iu/mL) | 47.5(32.5-121.8) | 50.6(35.75-507.9) | 0.740<sup>Z</sup> | 0.459 |
| TBG (ug/L) | 199.05(42-372.6) | 182.5(90.6-347.7) | 0.248<sup>Z</sup> | 0.805 |
| TRAb (iu/L) | 0.4(0.3-0.6) | 0.3(0.3-0.7) | -0.787<sup>Z</sup> | 0.431 |

h=hour, RAIU=radioiodine uptake, Hb=hemoglobin, TPOAb=anti-thyroid peroxidase
antibody, TGAb=anti-thyroglobulin antibody, TBG=thyroid binding globulin, TRAb=
anti-TSH receptor antibody.
<sup>Z</sup>Z-value of Wilcoxon rank test for two independent samples, <sup>T</sup>T-value of two independent536 samples t-test, <sup>F</sup>P-value of the fisher's exact test.

### Primary endpoints

As the primary endpoints, 18 patients in each group obtained effective outcomes from the treatments. Efficacy rates and recurrence rates were not statistically significantly different (Fig. 3). The ESR and CRP levels were statistically similar in the two groups at the time of drug withdrawal (Table 2).

**Table 2.** Comparisons for ESR and CRP between the two groups

| Characteristic | Time frame | Short-term<br>treatment<br>(n=26) | 6-weeks<br>treatment<br>(n=24) | Statistical<br>magnitude | P-value |
| --- | --- | --- | --- | --- | --- |
| ESR (mm/H) | Baseline | 65.5 (44-81) | 66 (41.5-73.5) | 0.29<sup>F1</sup> | 0.596 |
|  | 1 week | 15.5 (11-34) | 17.5 (10-42) | 122.47<sup>F2</sup> | <0.001 |
|  | Withdrawal | 15 (10-24) | 9 (6-17) | 1.59<sup>F3</sup> | 0.215 |
| CRP (mg/L) | Baseline | 27 (15-65) | 26 (15.5-42.5) | 0.01<sup>F1</sup> | 0.984 |
|  | 1 week | 5 (5-5) | 5 (5-5) | 18.68<sup>F2</sup> | <0.001 |
|  | Withdrawal | 5 (5-6.5) | 5 (5-10.2) | 0.29<sup>F3</sup> | 0.748 |

ESR=erythrocyte sedimentation rate, CRP=c-reactive protein.
<sup>F1</sup>F-value group effects of generalized linear mixed model (GLMM), <sup>F2</sup>F-value time effects of GLMM, <sup>F3</sup>F-value group \* time effects of GLMM.

**Figure 3.**
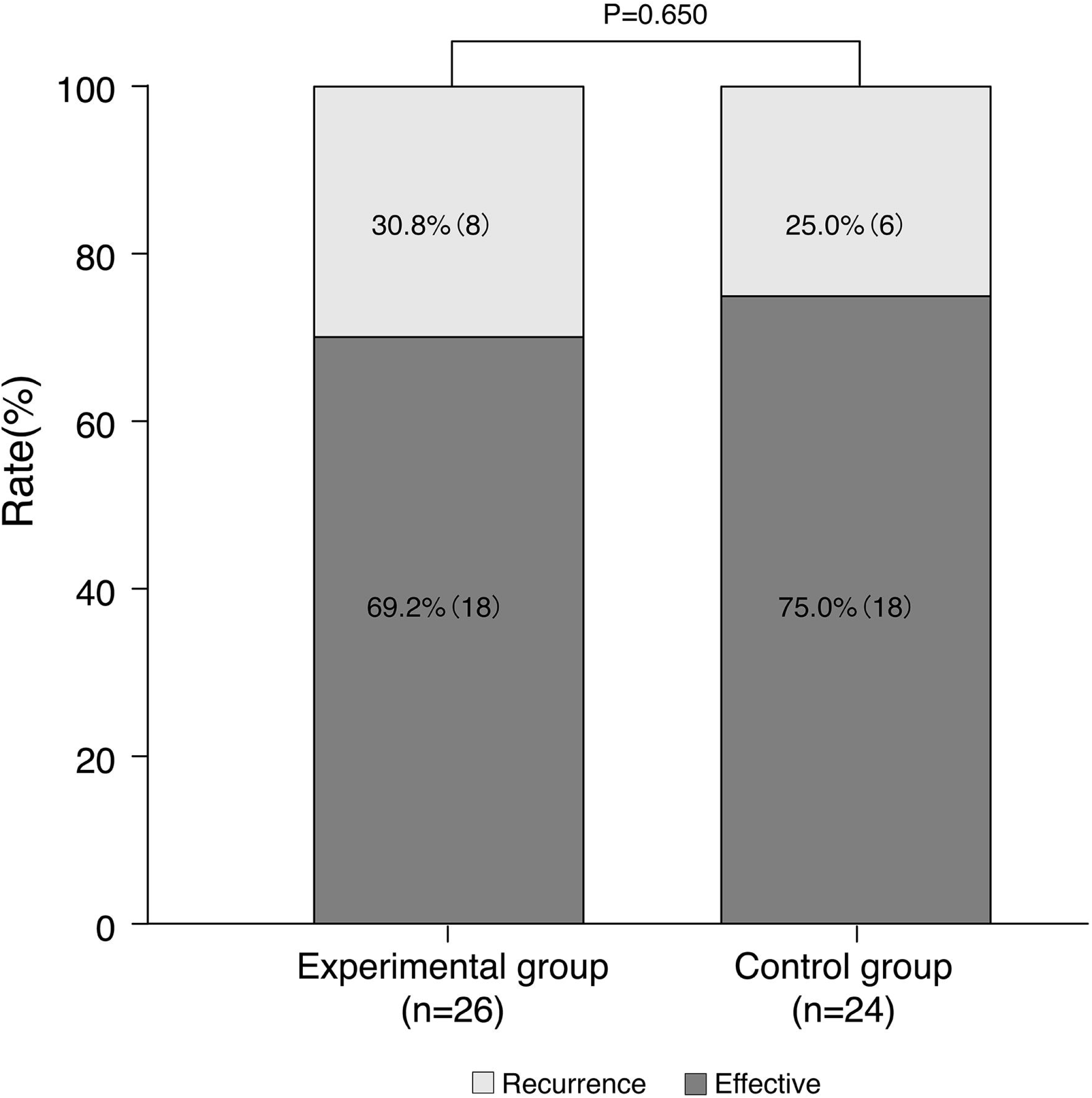
The effective and recurrent rates between experimental and control groups.

### Secondary endpoints

Regarding the secondary endpoints, PTH and SBP were significantly different between the two groups at the end of treatment. The PTH (28.8 vs 38.9 pg/ml, p=0.011) and mean SBP (113.9 vs 122.4 mmHg, p=0.023) at the time of withdrawal were significantly lower in the experimental group than in the control group.

No differences were observed in FT3, FT4, TSH, cortisol (Table 3), fasting plasma glucose (FPG), breakfast postprandial plasma glucose (BPPG), pre-dinner plasma glucose (PDPG), bedtime plasma glucose (BPG), GA, BCTX, OC, CT, 25-OH-VD, PINP, and TG, TC at baseline (Supplementary table). A difference was observed for baseline DBP, which ultimately did not influence baseline blood pressure or any primary and secondary endpoints.

**Table 3.**
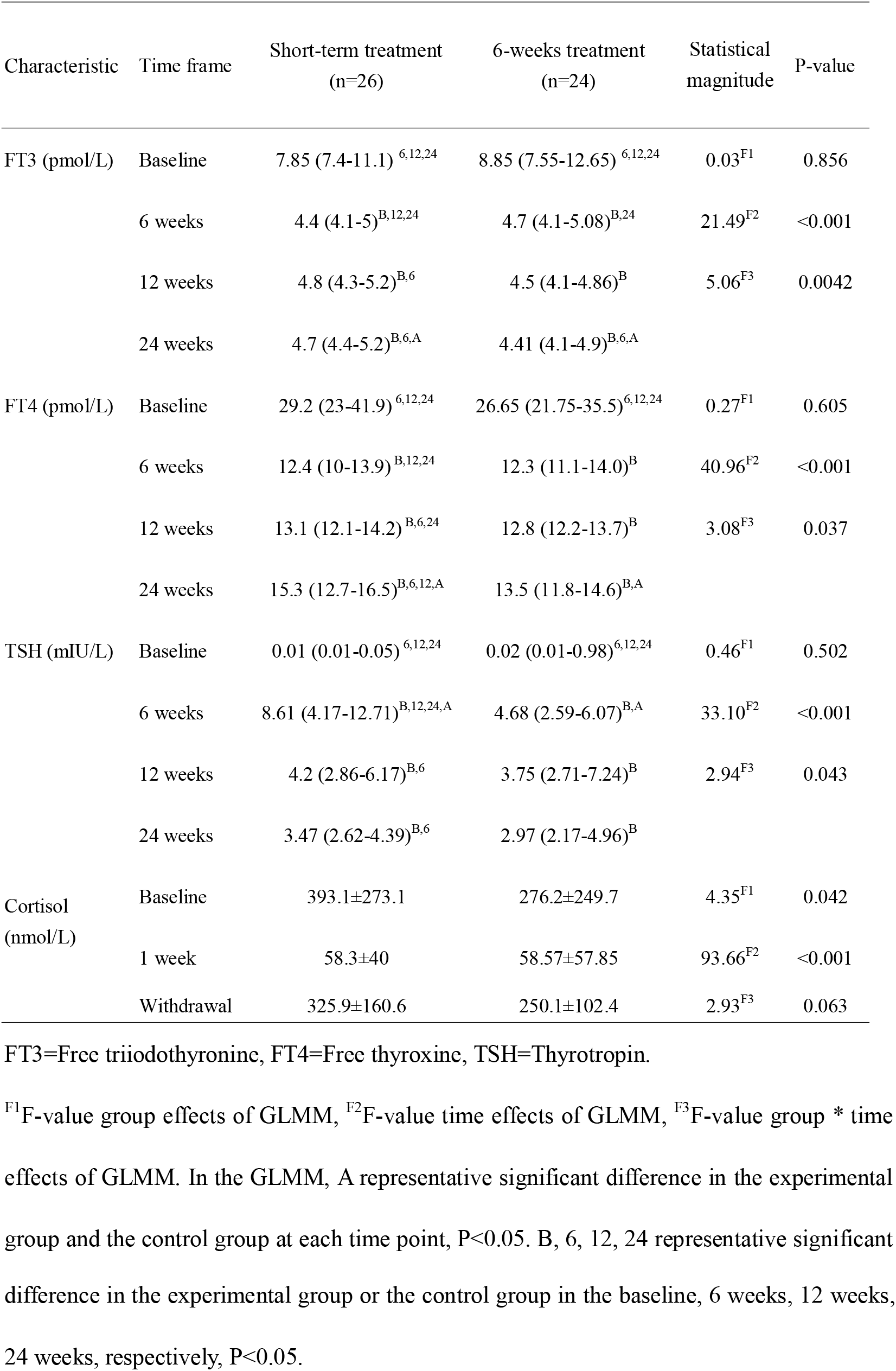
Comparisons for thyroid function and cortisol between the two groups

| Characteristic | Time frame | Short-term treatment<br>(n=26) | 6-weeks treatment<br>(n=24) | Statistical<br>magnitude | P-value |
| --- | --- | --- | --- | --- | --- |
| FT3 (pmol/L) | Baseline | 7.85 (7.4-11.1)<sup>6,12,24</sup> | 8.85 (7.55-12.65)<sup>6,12,24</sup> | 0.03<sup>F1</sup> | 0.856 |
|  | 6 weeks | 4.4 (4.1-5)<sup>B,12,24</sup> | 4.7 (4.1-5.08)<sup>B,24</sup> | 21.49<sup>F2</sup> | <0.001 |
|  | 12 weeks | 4.8 (4.3-5.2)<sup>B,6</sup> | 4.5 (4.1-4.86)<sup>B</sup> | 5.06<sup>F3</sup> | 0.0042 |
|  | 24 weeks | 4.7 (4.4-5.2)<sup>B,6,A</sup> | 4.41 (4.1-4.9)<sup>B,6,A</sup> |  |  |
| FT4 (pmol/L) | Baseline | 29.2 (23-41.9)<sup>6,12,24</sup> | 26.65 (21.75-35.5)<sup>6,12,24</sup> | 0.27<sup>F1</sup> | 0.605 |
|  | 6 weeks | 12.4 (10-13.9)<sup>B,12,24</sup> | 12.3 (11.1-14.0)<sup>B</sup> | 40.96<sup>F2</sup> | <0.001 |
|  | 12 weeks | 13.1 (12.1-14.2)<sup>B,6,24</sup> | 12.8 (12.2-13.7)<sup>B</sup> | 3.08<sup>F3</sup> | 0.037 |
|  | 24 weeks | 15.3 (12.7-16.5)<sup>B,6,12,A</sup> | 13.5 (11.8-14.6)<sup>B,A</sup> |  |  |
| TSH (mIU/L) | Baseline | 0.01 (0.01-0.05)<sup>6,12,24</sup> | 0.02 (0.01-0.98)<sup>6,12,24</sup> | 0.46<sup>F1</sup> | 0.502 |
|  | 6 weeks | 8.61 (4.17-12.71)<sup>B,12,24,A</sup> | 4.68 (2.59-6.07)<sup>B,A</sup> | 33.10<sup>F2</sup> | <0.001 |
|  | 12 weeks | 4.2 (2.86-6.17)<sup>B,6</sup> | 3.75 (2.71-7.24)<sup>B</sup> | 2.94<sup>F3</sup> | 0.043 |
|  | 24 weeks | 3.47 (2.62-4.39)<sup>B,6</sup> | 2.97 (2.17-4.96)<sup>B</sup> |  |  |
| Cortisol<br>(nmol/L) | Baseline | 393.1±273.1 | 276.2±249.7 | 4.35<sup>F1</sup> | 0.042 |
|  | 1 week | 58.3±40 | 58.57±57.85 | 93.66<sup>F2</sup> | <0.001 |
|  | Withdrawal | 325.9±160.6 | 250.1±102.4 | 2.93<sup>F3</sup> | 0.063 |

FT3=Free triiodothyronine, FT4=Free thyroxine, TSH=Thyrotropin.
<sup>F1</sup>F-value group effects of GLMM, <sup>F2</sup>F-value time effects of GLMM, <sup>F3</sup>F-value group \* time effects of GLMM. In the GLMM, A representative significant difference in the experimental group and the control group at each time point, P<0.05. B, 6, 12, 24 representative significant
difference in the experimental group or the control group in the baseline, 6 weeks, 12 weeks, 24 weeks, respectively, $P<0.05$ .

The FT3 and FT4 levels in both groups were significantly reduced compared to those at baseline at the 6th, 12th, 24th week, and resumed to normal reference range after 6 weeks. At the 6th week, the median of FT3 (normal range 3.1-6.8 pmol/L) decreased from 7.85 to 4.4 pmol/L in the experimental group and from 8.85 to 4.7 pmol/L in the control group. The median of FT4 (normal range 11-22 pmol/L) decreased from 29.2 to 12.4 pmol/L in the experimental group and from 26.65 to 12.3 pmol/L in the control group. The TSH levels of each group were significantly increased compared to baseline at the 6th, 12th, and 24th week (p<0.05 for all). At the 6th week TSH levels (normal range 0.27-4.20 mIU/L) increased from 0.01 to 8.61 mIU/L in the experimental group and from 0.02 to 4.68 mIU/L in the control group, both exceeding the upper limit of normal values, and fell to normal levels at the 12th and 24th week (Table 3). The incidences of transient overt and subclinical hyperthyroidism at diagnosis were 86% (43/50) (Fig. 4a), 81% (21/26), and 92% (22/24) in the general group, experimental group, and control group, respectively. The incidences of transient overt and subclinical hypothyroidism during follow-up period were 61% (30/49) (Fig. 4b), 72% (18/25), and 50% (12/24) in the general group, experimental group, and control group, respectively. Cortisol levels in the experimental and control groups were both significantly decreased to similar levels, from 393.1 and 276.2 mmol/L (baseline) to 58.3 and 58.57 mmol/L after 1 week of treatment (Table 3), respectively.

**Figure 4.**
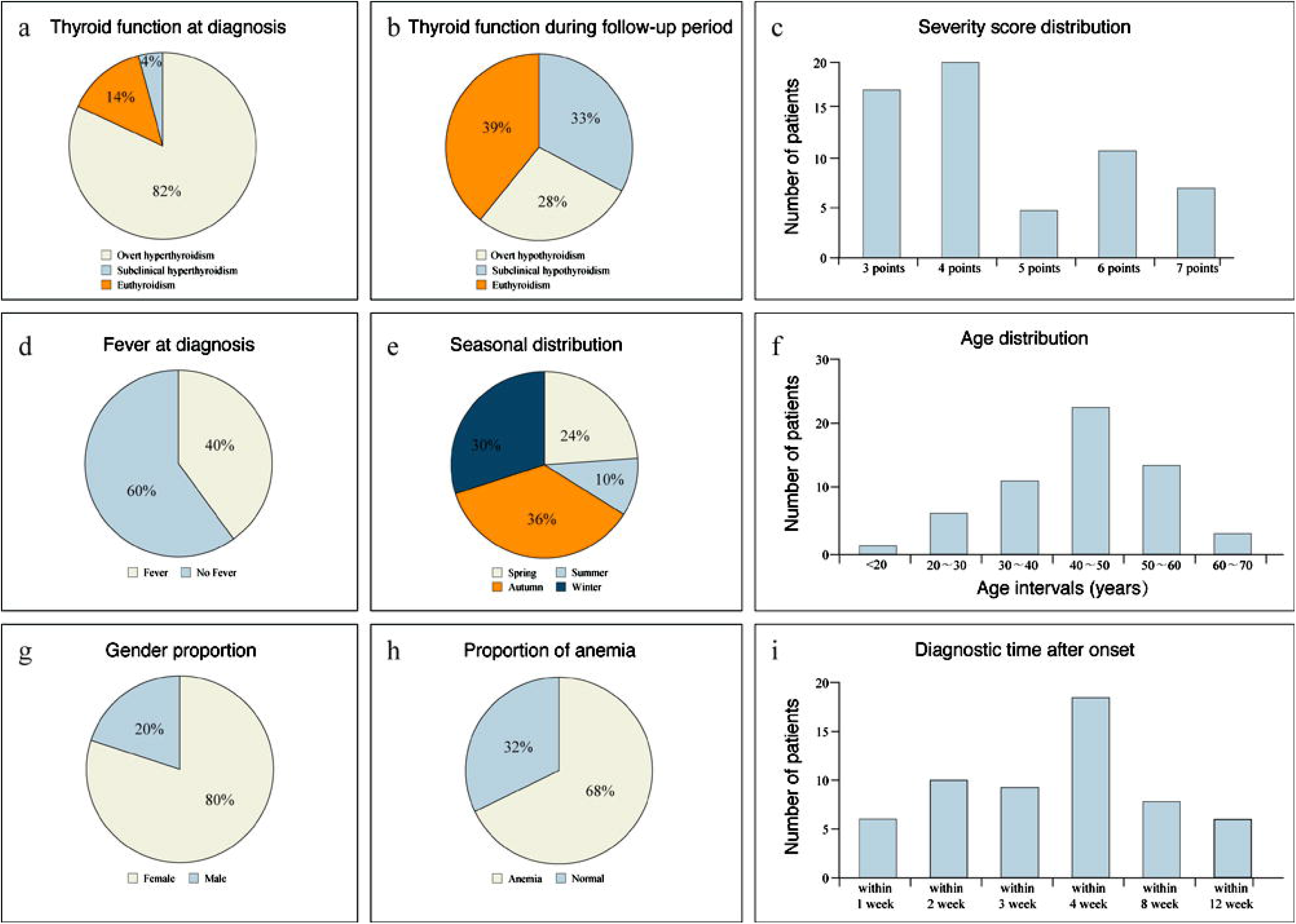
Other epidemiological data. a The incidence of subclinical hyperthyroidism, overt hyperthyroidism, and euthyroidism in the general group at diagnosis b The incidences of subclinical hypothyroidism, overt hypothyroidism, and euthyroidism in the general group during the 24-weeks follow-up period c The severity score distribution was based on the symptoms of SAT d The fever condition in the general group at diagnosis e The seasonal distribution in the general group at diagnosis f The age distribution in the general group at diagnosis g The gender proportion in the general group at diagnosis h The proportion of anemia in the general group at diagnosis i The diagnostic time after onset of this research

Compared with baseline levels, the mean FPG, BPPG, and GA levels were significantly decreased at the time of drug withdrawal in both groups. Mean FPG reduced by 0.9 mmol/L in the experimental group and 0.61 mmol/L in the control group. Mean BPPG reduced by 1.09 mmol/L in the experimental group and 0.89 mmol/L in the control group. Mean GA levels reduced by 1.81% in the experimental group and 2.06% in the control group. Furthermore, the mean PDPG of the control group was significantly decreased at the time of drug withdrawal compared with baseline (Supplementary table 1).

The median PINP in the experimental group (increased to 47.14 ng/mL) and in the control group (increased to 45.46 ng/mL) was significantly elevated at the time of withdrawal compared with baseline. The mean OC level in the control group (increased to 23.1 ng/mL) was significantly increased at the time of drug withdrawal compared to baseline. TG and TC levels of the two treatment groups were significantly increased at the time of drug withdrawal compared to baseline (TG increased by 0.44 mmol/L in the experimental group and 0.52 mmol/L in the control group, TC increased by 1.82 mmol/L in the experimental group and 1.42 mmol/L in the control group) (Supplementary table 2).

A total of 14 patients developed recurrence and received the new treatment method. Withdrawal occurred in 92.9% of all relapsed patients after 4 weeks of therapy, except one patient whose treatment was discontinued at the 7^th^ week.

## Discussion

We designed a randomized controlled trial to evaluate feasibility of short-term prednisone as a strategy for treating moderate-to-severe symptoms of SAT. Furthermore, our goal was to minimize the side effects of glucocorticoids while maintaining efficacy. We therefore administered prednisone for 1 week and then switched to NSAIDs for 1 week in patients in the experimental group. During the 24-week follow-up period, the experimental group showed non-inferior efficacy compared with the control group, with fewer side effects.

Owing to the wide-range of suppressive effects on chemical mediators [18], patients receiving corticosteroids for SAT had a shorter time period to symptom resolution compared with those treated with NSAIDs [19]. Therefore, the guidelines recommend that patients failing to respond to NSAIDs or those with moderate-to-severe symptoms should be treated with corticosteroids such as prednisone 40 mg daily for 1-2 weeks [1]. Doses should be gradually tapered over 2-4 or 4-6 weeks after relief of symptoms [20]. However, the slow reduction results in a long course of treatment, more than 8 weeks in some cases [3]. One study recommended corticosteroid withdrawal until iodine uptake resumed to normal. The course of treatment was at least 76 days [17]. Even short-term glucocorticoid use for less than 30 days increased the incidence of sepsis, venous thromboembolism, and fracture [21]. Thus, managing the side effects of glucocorticoids is challenging. The ideal treatment options include minimized effective doses or keeping the course as short as possible [3]. At present, RCTs examining shorter courses of prednisone are lacking. In general, patients given acute corticosteroid therapy (< 7-14 days) do not develop HPA axis suppression. As a result, treatment can be suspended [22]. Similar to another study [12], the ESR was still elevated after 1 week administration of 30 mg prednisone daily in our preliminary experiment. Additionally, NSAIDs are considered as a first-line therapy to relieve pain in SAT with mild symptoms. Subsequently, we added 1 week of celecoxib in the short-term group, with the aim to eliminate SAT completely. Therefore, the RCT was conducted using 1 week of prednisone, then 1 week of NSAIDs, compared with the 6-week regimen recommended by the Chinese guidelines.

Our research demonstrated that the experimental and control groups had similar efficacy and recurrence rates, consistent with previous studies [12,13,16]. Recurrence is frequent and can occur at any time during or after treatment [11]. In the experimental group, 3 patients relapsed during the treatment period, and 5 relapsed after withdrawal. In the control group, recurrence occurred in 2 patients during the treatment period and 4 patients after withdrawal. These recurrence characteristics reminded us that the relapse was possible after withdrawal. As the dosage was decreased, approximately 20% of patients relapsed and required restoration of the higher dose [16]. In our research, treatment using 10 mg prednisone in the morning and 10 mg in the afternoon was used for relapse management.

At present, there is no one systematic review to assess mild and overlooked influences of glucocorticoid, such as on blood glucose and blood pressure, during SAT treatment [3]. We first systematically observed these side effects. At the end of treatment, SBP was lower (by approximately 10 mmHg) in the experimental group than in the control group, which indicated that short-term prednisone might have less effect on SBP. Increased plasma volume, elevated peripheral vascular resistance and heightened cardiac output are the possible mechanisms contributing to the development of hypertension with glucocorticoid excess. Increased plasma volume may be related to long-term glucocorticoids acting on mineralocorticoid receptors [23]. We also found elevated PTH levels, even if prednisone was taken only for 1 week. Similarly, PTH was significantly lower (by approximately 10 pg/mL) in the experimental group than in the control group. Glucocorticoid inhibits calcium absorption in the small intestine by affecting vitamin D, thereby reducing serum calcium and slightly elevating PTH, which leads to secondary osteoporosis [24]. Moreover, PINP, the indirect marker of bone formation, was elevated at withdrawal compared with that at baseline in both groups, which indicated that prednisone could affect bone formation even if the course of treatment was only 1 week. Similar to a previous study [25], we observed that prednisone can significantly elevate blood lipids, such as TG and TC, even with short-term use. In addition, FPG, BPPG, and GA were all decreased, suggesting that the glucose-lowering effect of pain relief during stress may be stronger than the hyperglycemic effect that antagonizes insulin after prednisone use. Increased OC and decreased PDPG at withdrawal, compared with that at baseline in the control group, showed only a changing trend with no significant difference in the experimental group. These may reach statistical difference after expanding the sample size. The symptomology of chronic adrenal insufficiency was not observed, and there was no life-threatening acute adrenal crisis in the two groups.

Although many researchers deemed transient hyperthyroidism and hypothyroidism are universal after SAT, we determined permanent hypothyroidism may not be so common. In our study, the incidences of temporary hyperthyroidism and hypothyroidism were much higher than enrolled patients in other trials [1]. We speculated that the higher incidence compared to previous studies in our study which may have included more severe patients with thyroid follicle damage. These patients likely experienced transient hyperthyroidism and slow thyroid follicle repair, leading to lower thyroid hormones levels to stimulate pituitary TSH secretion. The incidence of persistent hypothyroidism is 5-15% [1], and patients with permanent hypothyroidism require life-long thyroxine therapy [16]. There were only 2 patients with negative thyroid autoantibodies who had “persistent hypothyroidism” during the 24-week follow-up period. However, levothyroxine dose gradually reduced until ultimate withdrawal along with the extension of follow-up period to 3 years 8 months and 4 years, respectively. The follow-up time in one study [26] was long enough to be similar to our findings, without occurrences of persistent hypothyroidism.

It’s our novelty that we introduced the scoring criteria [6] to calculate symptom severity scores of patients in this study (Fig. 4c). Two studies [27, 28] demonstrated fever development in 46.2% of patients (>38.3°C) and 28.2% (>38°C) of patients. In this study, 40% of patients had fever (>37°C), suggesting that fever was common in moderate-to-severe SAT (Fig. 4d). Our results also suggest that the 90% of SAT cases occurred in the spring, autumn and winter months in China (Fig. 4e). Different seasonal clusters during summer to early autumn [27] and in the late summer and autumn [29] have been reported in other countries. Consistent with the literature [30], the predilection is 86% in those aged 30-60 years and especially 42% in those aged 40-50 years (Fig. 4f). Similar to American communities [4], women were more susceptible to SAT, and the ratio of female to male cases was 4:1 (Fig. 4g). Mild anemia was common, with an incidence rate of 68% (Fig. 4h). Our average time to definite diagnosis was 4.46 weeks (Fig. 4i), suggesting that the disease was easily missed initially. When thyroid enlargement was assessed by palpation, the rate of normalization after 7 days of prednisone treatment was 100%. After 24 and 48 hours treatment, the pain relief rates were 90% and 98%, suggesting that prednisone is a particularly effective drug for moderate-to-severe SAT. Similar to several other reports [6, 11, 28], our positive rates of TPOAb and TGAb were 20.41% and 37.5%. TRAb is often significantly elevated in patients with Graves’ disease. In this research, the rate of TRAb exceeding the upper limit of normal was 2.33%, suggesting that TRAb is a valuable indicator for differentiating Graves’ disease from hyperthyroidism caused by SAT. TBG levels have been used as a marker for monitoring early-phase SAT [12]. The rate of TBG exceeding the normal value in our study was 72.73%, suggesting that thyroid follicular destruction and TBG release are common phenomena in SAT. Similar to other reports [29], there was a low RAIU, which could be used to distinguish this condition from Graves’ disease during the toxic phase of SAT.

The present study had several limitations. First, we did not analyze that the side effects of celecoxib, which may have affected the experimental results. Second, our study included a small sample size; thus, the results should be verified in a larger sample size. Third, this was a single-center and single-blinded study, which limits the strength of our findings. A double-blinded, multicenter study would be the next step.

## Conclusions

Overall, current treatment recommendations are based on observational data and expert opinion. This is the first randomized controlled trial to evaluate the efficacy and safety of 1-week vs 6-week treatment for moderate-to-severe SAT. Our study demonstrated that the two groups had similar rates of effectiveness and recurrence. Furthermore, the short-term treatment had fewer side effects with respect to bone metabolism and blood pressure. Short-term glucocorticoid with a better safety profile may be likely an optional strategy for moderate-to-severe SAT.

## Data Availability

The datasets generated and/or analysed during the current study are not publicly available due to individual privacy but are available from the corresponding author on reasonable request.

## Abbreviations

SAT: Subacute thyroiditis; ESR: Erythrocyte sedimentation rate; CRP: C-reactive protein; NSAIDs: Nonsteroidal anti-inflammatory drugs; RCTs: Randomized controlled trials; Hb: Hemoglobin; TSH: Thyroid-stimulating hormone; FT3: Free triiodothyronine; FT4: Free thyroxine; GA: Glycated albumin; TC: Total cholesterol; TG: Triglyceride; SBP: Systolic blood pressure; DBP: Diastolic blood pressure; BCTX: β cross-linked C-telopeptide of type 1 collagen; OC: osteocalcin; PINP: Type I procollagen amino terminal peptide; PTH: Parathyroid hormone; 25-OH-VD: 25-hydroxy vitamin D; CT: Calcitonin; TGAb: Anti-thyroglobulin antibody; TPOAb: Anti-thyroid peroxidase antibody; TBG: Thyroid-binding globulin; TRAb: Anti-TSH receptor antibody; RAIU: Radioiodine uptake; FPG: Fasting plasma glucose; BPPG: Breakfast postprandial plasma glucose; PDPG: Pre-dinner plasma glucose; BPG: Bedtime plasma glucose

## Declarations

### Ethics approval and consent to participate

This study was approved by the Ethics Committee of Xinqiao Hospital, Third Military Medical University, China (reference number 2018012-01).

### Consent for publication

Participants provided written informed consent for publication of research results.

## Authors’ contributions

LD was involved with the literature search, study design, clinical diagnosis, patient follow-up, data collection and writing of the first and revised draft of the manuscript. XF, RZ, XT, XX, SF and HZ were chief investigator. SF was involved with revision of the manuscript. All other authors critically reviewed and approved the final version.

## Acknowledgments

The authors would like to thank Yetao Luo and Yuan Liu for statistical analysis.

## References

1. Ross DS, Burch HB, Cooper DS, Greenlee MC, Laurberg P, Maia AL, Rivkees SA, Samuels M, Sosa JA, Stan MN,Walter MA. 2016 American Thyroid Association Guidelines for Diagnosis and Management of Hyperthyroidism and Other Causes of Thyrotoxicosis. Thyroid. 2016;26(10):1343–1421.

2. Bahn Chair RS, Burch HB, Cooper DS, Garber JR, Greenlee MC, Klein I, Laurberg P, McDougall IR, Montori VM, Rivkees SA,Ross DS,Sosa JA, Stan MN. Hyperthyroidism and Other Causes of Thyrotoxicosis: Management Guidelines of the American Thyroid Association and American Association of Clinical Endocrinologists. Thyroid. 2011;21(6):593–646.

3. Kubota S, Nishihara E, Kudo T, Ito M, Amino N, Miyauchi A. Initial Treatment with 15[mg of Prednisolone Daily Is Sufficient for Most Patients with Subacute Thyroiditis in Japan. Thyroid. 2013;23(3):269–272.

4. Oray M, Abu Samra K, Ebrahimiadib N, Meese H, Foster CS. Long-term side effects of glucocorticoids. Expert Opin Drug Saf. 2016;15(4):457–465.

5. Broersen LHA, Pereira AM, Jørgensen JOL, Dekkers OM. Adrenal Insufficiency in Corticosteroids Use: Systematic Review and Meta-Analysis. J Clin Endocrinol Metab. 2015;100(6):2171–2180.

6. Benbassat CA, Olchovsky D, Tsvetov G, Shimon I. Subacute thyroiditis: Clinical characteristics and treatment outcome in fifty-six consecutive patients diagnosed between 1999 and 2005. J Endocrinol Invest. 2007;30(8):631–635.

7. Koirala KP, Sharma V. Treatment of Acute Painful Thyroiditis with Low Dose Prednisolone: A Study on Patients from Western Nepal. J Clin Diagn Res. 2015;9(9): MC01–03

8. Arao T, Okada Y, Torimoto K, Kurozumi A, Narisawa M, Yamamoto S, Tanaka Y. Prednisolone Dosing Regimen for Treatment of Subacute Thyroiditis. J UOEH. 2015;37(2):103–10.

9. Floriani C, Feller M, Aubert CE, M’Rabet-Bensalah K, Collet TH, den Elzen WPJ, Bauer DC, Angelillo-Scherrer A, Aujesky D, Rodondi N. Thyroid Dysfunction and Anemia: A Prospective Cohort Study and a Systematic Review. Thyroid. 2018;28(5):575–582.

10. Garber JR, Cobin RH, Gharib H, Hennessey JV, Klein I, Mechanick JI, Pessah-Pollack R, Singer PA, Woeber KA. Clinical practice guidelines for hypothyroidism in adults: cosponsored by the American Association of Clinical Endocrinologists and the American Thyroid Association. Thyroid. 2012;22(12):1200–1235.

11. Fatourechi V, Aniszewski JP, Fatourechi GZE, Atkinson EJ, Jacobsen SJ. Clinical Features and Outcome of Subacute Thyroiditis in an Incidence Cohort: Olmsted County, Minnesota, Study. J Clin Endocrinol Metab. 2003;88(5):2100–2105.

12. Mizukoshi T, Noguchi S, Murakami T, Futata T, Yamashita H. Evaluation of recurrence in 36 subacute thyroiditis patients managed with prednisolone. Intern Med. 2001;40(4):292–295.

13. Bennedbaek FN, Hegedus L. The value of ultrasonography in the diagnosis and follow-up of subacute thyroiditis. Thyroid. 1997;7(1):45–50.

14. Topuzovic N, Smoje J, Karner I. The therapeutic approach in subacute (de Quervain’s) thyroiditis. J Nucl Med. 1997;38(10):1665.

15. Yamada T, Sato A, Aizawa T. Dissociation between serum interleukin-6 rise and other parameters of disease activity in subacute thyroiditis during treatment with corticosteroid. J Clin Endocrinol Metab. 1996;81(2):577–579.

16. Volpe R. The management of subacute (DeQuervain’s) thyroiditis. Thyroid. 1993;3(3):253–255.

17. Vagenakis AG, Abreau CM, Braverman LE. Prevention of recurrence in acute thyoiditis following corticosteroid withdrawal. J Clin Endocrinol Metab. 1970;31(6):705–708.

18. Cunningham FM, Lees P. Advances in anti-inflammatory therapy. Br Vet J. 1994;150(2):115–134.

19. Sato J, Uchida T, Komiya K, Goto H, Takeno K, Suzuki R, Honda A, Himuro M, Watada H. Comparison of the therapeutic effects of prednisolone and nonsteroidal anti-inflammatory drugs in patients with subacute thyroiditis. Endocrine. 2017;55(1):209–214.

20. Pearce EN, Farwell AP, Braverman LE. Thyroiditis. N Engl J Med. 2003;348(26):2646–2655.

21. Waljee AK, Rogers MAM, Lin P, Singal AG, Stein JD, Marks RM, Ayanian JZ, Nallamothu BK. Short term use of oral corticosteroids and related harms among adults in the United States: population based cohort study. BMJ. 2017:357:j1415.

22. Alves CE, Sio, Robazzi TCV, Ccedil M, A M. Withdrawal from glucocorticosteroid therapy: clinical practice recommendations. J Pediatr (Rio J). 2008;84(3):192–202.

23. Pimenta E, Wolley M, Stowasser M. Adverse Cardiovascular Outcomes of Corticosteroid Excess. Endocrinology. 2012;153(11):5137–5142.

24. Mirza F, Canalis E. Management of endocrine disease: Secondary osteoporosis: pathophysiology and management. Eur J Endocrinol. 2015;173(3):R131–R151.

25. Taskinen MR, Kuusi T, Yki-Jarvinen H, Nikkila EA. Short-term effects of prednisone on serum lipids and high density lipoprotein subfractions in normolipidemic healthy men. J Clin Endocrinol Metab. 1988;67(2):291–299.

26. Alfadda AA, Sallam RM, Elawad GE, AlDhukair H, Alyahya MM. Subacute Thyroiditis: Clinical Presentation and Long Term Outcome. Int J Endocrinol. 2014;2014:794943.

27. Nishihara E, Ohye H, Amino N, Takata K, Arishima T, Kudo T, Ito M, Kubota S, Fukata S, Miyauchi A. Clinical characteristics of 852 patients with subacute thyroiditis before treatment. Intern Med. 2008;47(8):725–729.

28. Erdem N, Erdogan M, Ozbek M, Karadeniz M, Cetinkalp S, Ozgen AG, Saygili F, Yilmaz C, Tuzun M, Kabalak T. Demographic and clinical features of patients with subacute thyroiditis: results of 169 patients from a single university center in Turkey. J Endocrinol Invest. 2007;30(7):546–550.

29. Sweeney LBM, Stewart CM, Gaitonde DYM. Thyroiditis: An Integrated Approach. Am Fam Physician. 2014;90(6):389–396.

30. Singer PA. Thyroiditis. Acute, subacute, and chronic. Med Clin North Am. 1991;75(1):61–77.

